# Physicians in the management and leadership of health care: A systematic review of the conditions conducive to organizational performance

**DOI:** 10.1101/19011379

**Authors:** Mairi Savage, Carl Savage, Mats Brommels, Pamela Mazzocato

## Abstract

**Introduction:** The influx of management ideas into health care has triggered considerable debate about if and how managerial and medical logics can co-exist. Recent reviews suggest that clinician involvement in hospital leadership can lead to superior performance.

**Objective:** To systematically explore the conditions instrumental for medical leadership to have an impact on organizational performance.

**Methods:** We searched PubMed, Web of Science, and Psychinfo for peer-reviewed, empirical, English language articles and reviews published between January 1, 2006 and August 12, 2018. We performed a thematic synthesis through inductive line-by-line coding of the included studies.

**Results:** The search yielded 1447 publications, of which 62 were included. Three major themes were identified that described a movement 1. From medical protectionism to management through medicine, 2. From command and control to participatory leadership practices, and 3. Organizational practices to support incidental versus willing leaders. Based on these themes, the authors developed a model to depict conditions that facilitate or impede the influence of medical leadership through a virtuous cycle of management through medicine or a vicious cycle of medical protectionism.

**Conclusions:** This review helps individuals, organizations, educators, and trainers better understand how medical leadership can be both a boon and a barrier to the performance of health care organizations. In contrast to the conventional view of conflicting logics, medical leadership would benefit from a more integrative mental model of management and medicine. Nurturing medical engagement requires participatory leadership enabled through long-term investments at the individual, organizational, and system levels. These combined efforts will enable a shift to new leadership paradigms suitable to the complexity of health care, and establish conditions favorable for large-system transformation and health care reform.

**Strengths and limitations of this study:**

- While previous literature reviews have established a correlation between physicians in leadership roles and organizational performance, this is to our knowledge the first review that seeks to explore what contributes to that link.
- The review expands on the typically quantitative focus of systematic reviews by providing a thematic synthesis of fifty-five empirical studies and seven literature reviews.
- A model is presented that depicts a virtuous cycle of management through medicine and a vicious cycle of medical protectionism.
- This review is limited by the quality and heterogeneity of the included studies.
- While plausible correlations between conditions and performance outcomes are explored, to establish causality requires study designs that can determine the strength of the relationships.

## INTRODUCTION

Organizational research has established a link between leadership practices and performance.[1] As health care searches for its success formula, the impact of medical leadership on performance has become an increasingly relevant research objective. The two most recent systematic reviews on the subject suggest that clinician involvement in hospital leadership can be linked to superior performance.[2,3] The inclusion of clinical leaders (primarily physicians) in senior management roles has a positive impact on care quality, management of financial and operational resources, and social performance, albeit a few studies showed a negative impact on the latter two.[2] Additional reviews have found effects on staff satisfaction, retention, performance, and burnout;[4–6] psychological safety, respect, and shared goals;[7] approval and support of political reforms[8]; and the adoption of information technology.[9]

While the reviews describe the challenge to discern why medical leadership makes a difference, Sarto and Veronesi,[2] hypothesize about possible mediating mechanisms (Figure 1).

**Figure 1.**
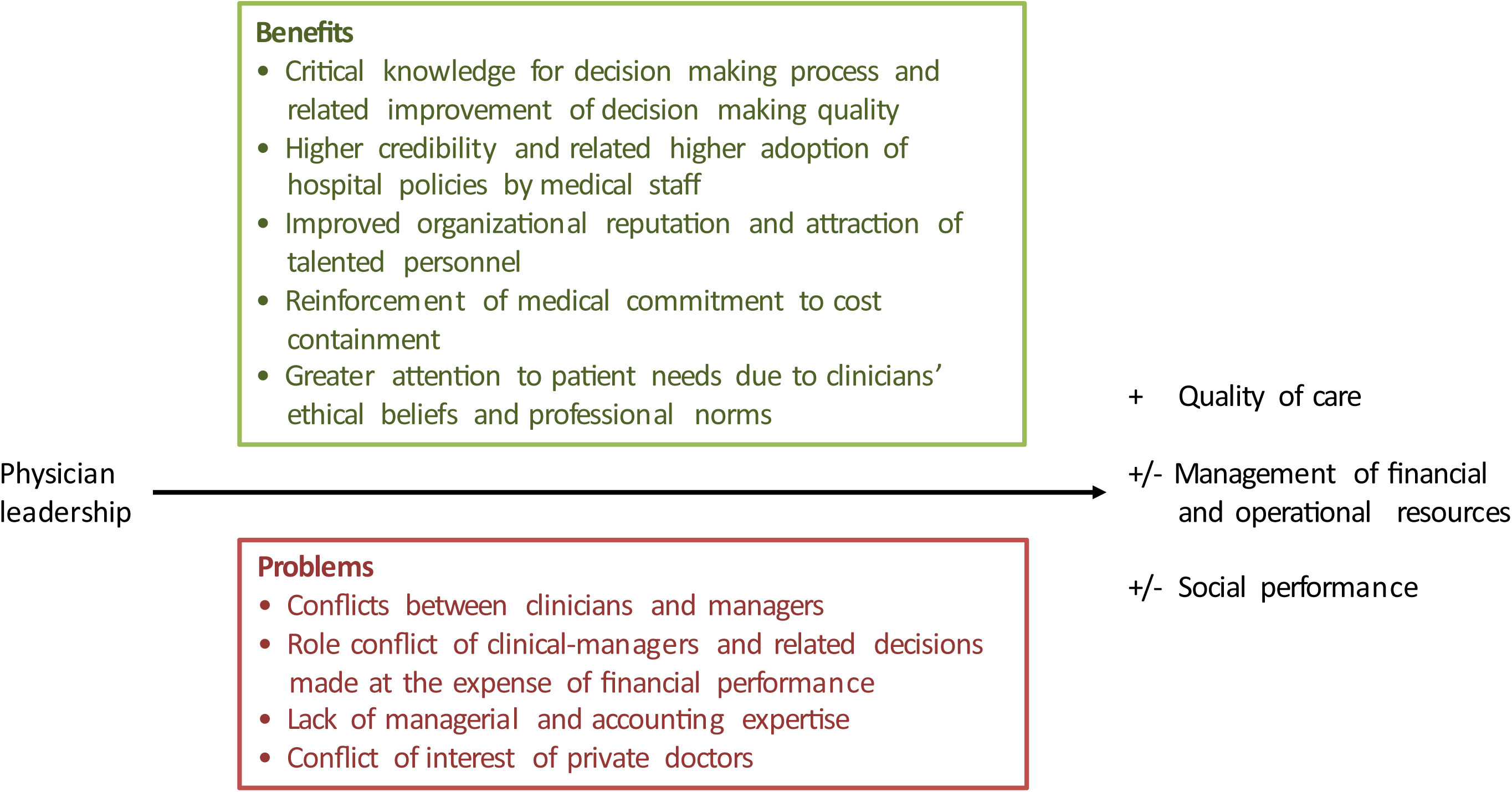
An explanatory model of factors that mediate the positive and negative effects of physician leadership (adapted from (Sarto and Veronesi 2016)).

The core explanation proffered is centered on the individual’s credibility and competence generated by a medical degree.[2] However, two observations can be made, both of which warrant a further qualitative exploration. The first is that the mediating mechanisms are drawn from authors discussions of their quantitative results rather than research designed to specifically explore the mechanisms behind the connections. The second is that the mediating mechanisms exist within a context,[10] i.e. there are conditions that influence to what extent medical competence and credibility can benefit organizational performance. The aim of this study is therefore to systematically explore the conditions instrumental for medical leadership to have an impact on organizational performance.

## METHODS

### Review protocol

This systematic literature review is a thematic synthesis of empirical studies and literature reviews. Thematic synthesis was chosen in order to expand the traditionally quantitative focus of systematic reviews with a method that accommodates a diversity of study designs, provides policy-makers and practitioners more nuanced evidence for a complex question,[11] and enables to develop insights beyond those of the original studies through an higher-order thematic structure.[11,12] Given its qualitative nature, it was guided by the ENhancing Transparency in REporting the synthesis of Qualitative research (ENTREQ) statement (Appendix 1).[13] Patients or the public were not involved in the design, or conduct, or reporting, or dissemination plans of our research.

### Search strategy

The strategy was developed with assistance from a professional research librarian. We conducted a comprehensive search for scientific articles published between January 1^st^ 2006 and August 12^th^ 2018. We limited the search timeline to capture contemporary evidence in the light of recently established correlations between medical leadership and performance.[2] We defined this as the last decade of publications. As the study originally commenced in 2016, we updated the search on 12^th^ of August 2018. Boolean searches were performed in Medline/PubMed, Web of Science, and Psychinfo. As the focus was on physicians, other health care databases such as CINAHL, were excluded. To identify a wide range of studies, all possible truncated combinations of keywords and MeSH terms such as “clinical/medical/physician/doctor management/leadership”, “organization and management”, “physician executive”, “performance”, and “quality of health care” were used. The search was complemented with additional articles from the reference lists of the articles selected for full-text review.

### Study selection

Aggregated search results were imported to the Mendeley reference management system where duplicates were removed. Remaining records were subjected to three rounds of screening. Inclusion criteria were that articles were peer-reviewed, empirical studies or literature reviews, and in the English language, published between January 2006 and August 2018 which focused on physicians in the leadership and management of health care. We included literature reviews to capture patterns across a wide span of studies, i.e. we did not use these to assess the relative importance of individual factors, but rather to identify relevant themes in the literature.

Exclusion criteria were publication prior to 2006, non-English language, not empirical or literature reviews, non-peer-reviewed, did not include physicians as study participants, and were reports on care and treatment planning for specific medical conditions. These inclusion and exclusion criteria were applied, when the first author first screened all titles and key words, and then the remaining abstracts. Then, all authors screened the records eligible for full text review and applied further exclusion criteria: full-text not available; purely quantitative reports on organizational performance outcomes or leadership development evaluations; not addressing physicians in the leadership and management of health care (i.e. not about their role in quality improvement, coordination of care, resource management, team leadership, change management, policy reform, or descriptions of their individual experiences in such roles). Any discrepancies regarding inclusion were resolved through consensus. Due to the diversity of study designs and contexts, and the intention to capture a thematic account, the quality of individual studies in terms of strength of evidence was not assessed as per established convention.[13,14]

### Data extraction and analysis

Data on general characteristics included type of study design, country of origin, setting, and study participants. Data extraction and analysis followed an inductive approach. The results sections were read line-by-line to identify meaning units describing the conditions instrumental to medical leadership. The first author summarized these as codes, which were then organized into descriptive themes by all authors.[12] Based on these themes, the authors developed a preliminary model (analytical themes) to depict conditions that facilitate or impede physician leadership.[12] The model was tested for face validity and refined to improve clarity after discussions with practicing clinicians and managers in our graduate and continuing professional development courses and at conferences in Sweden and Europe. Data extraction and analysis was performed in NVivo qualitative data analysis software; QSR International Pty Ltd. Version 10, 2012.

## RESULTS

The initial search identified 1447 records (PubMed 437, Web of Science 896, and Psychinfo 114). After removing duplicates and adding 26 records identified from reference lists, the tally was 1424 records. Titles and key words were screened which yielded 367 records. After their abstracts were screened,189 articles remained. After screening the full texts, 62 articles were included in the thematic synthesis (Figure 2). Of these, fifty-five were empirical articles (qualitative, quantitative or mixed methods designs) and seven literature reviews.

**Figure 2.**
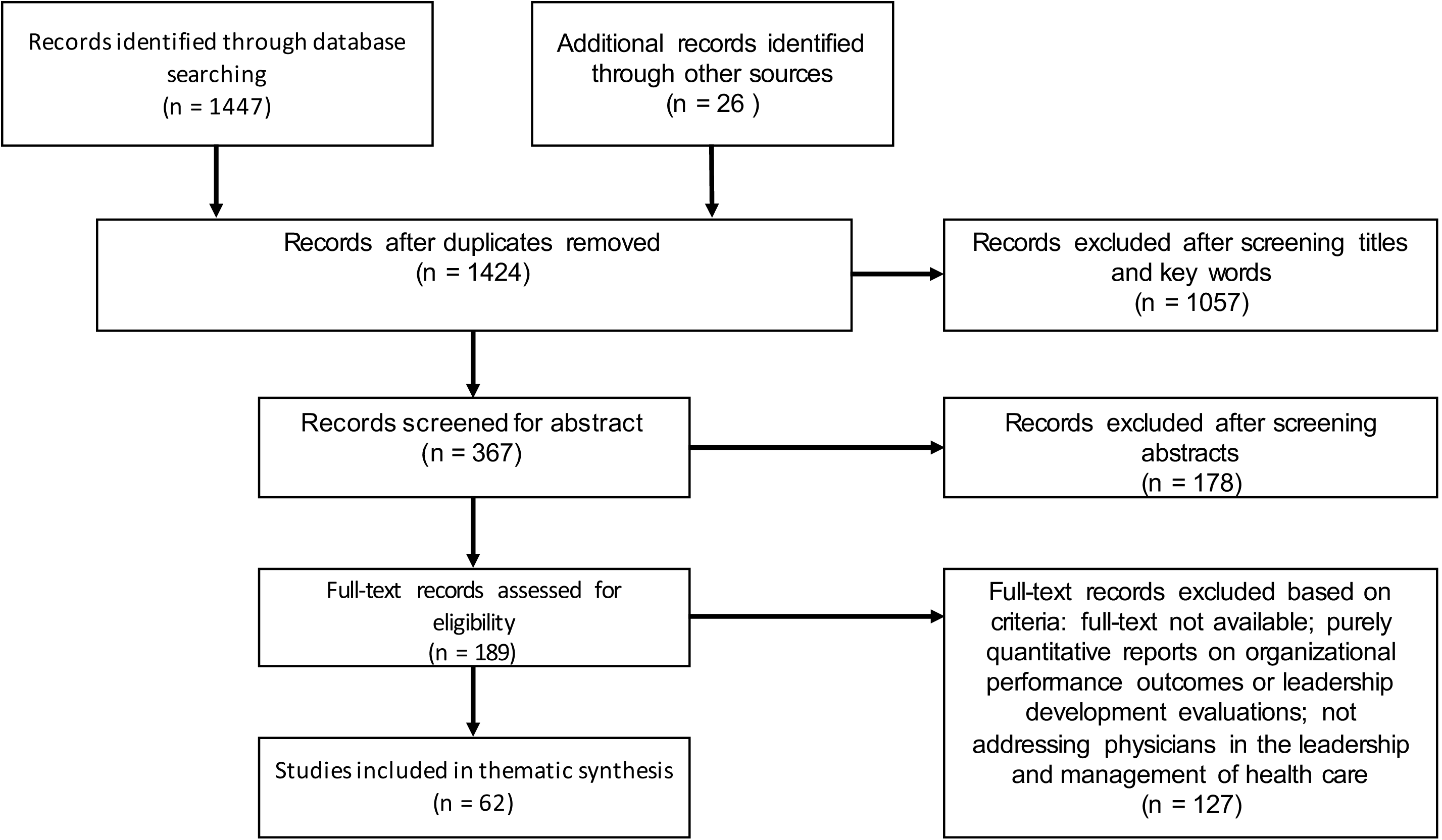
PRISMA Flowchart.

### General characteristics

Most studies were conducted in the UK (n=17) and the US (n=14), in hospital settings (n=40), and focused on senior managers (n=14). Qualitative designs were used in 23 studies, followed by 12 surveys and 10 case studies (Figure 3). The empirical studies together reported on 906 hours of observations, 1417 interviews, and 22643 survey responses. A detailed overview of the included studies is provided in Appendix 2.

**Figure 3.**
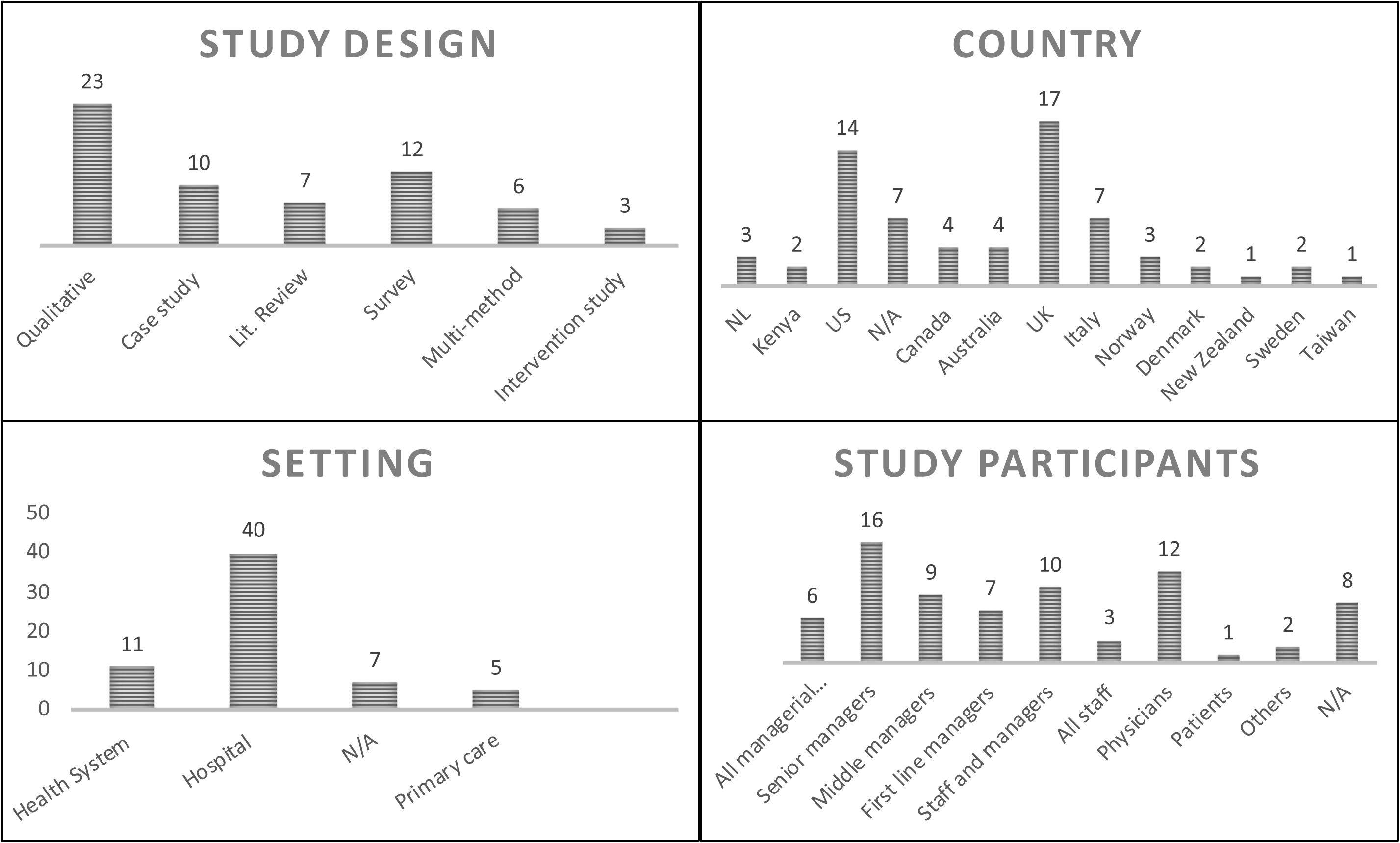
General characteristics of included studies.

### Conditions instrumental for medical leadership to have an impact on organizational performance

Three themes were identified: From medical protectionism to management through medicine; from command and control to participatory leadership practices; and organizational practices to nurture willing *vs*. incidental leaders (Table 1). References to the relevant articles are provided in the text.

**Table 1.**
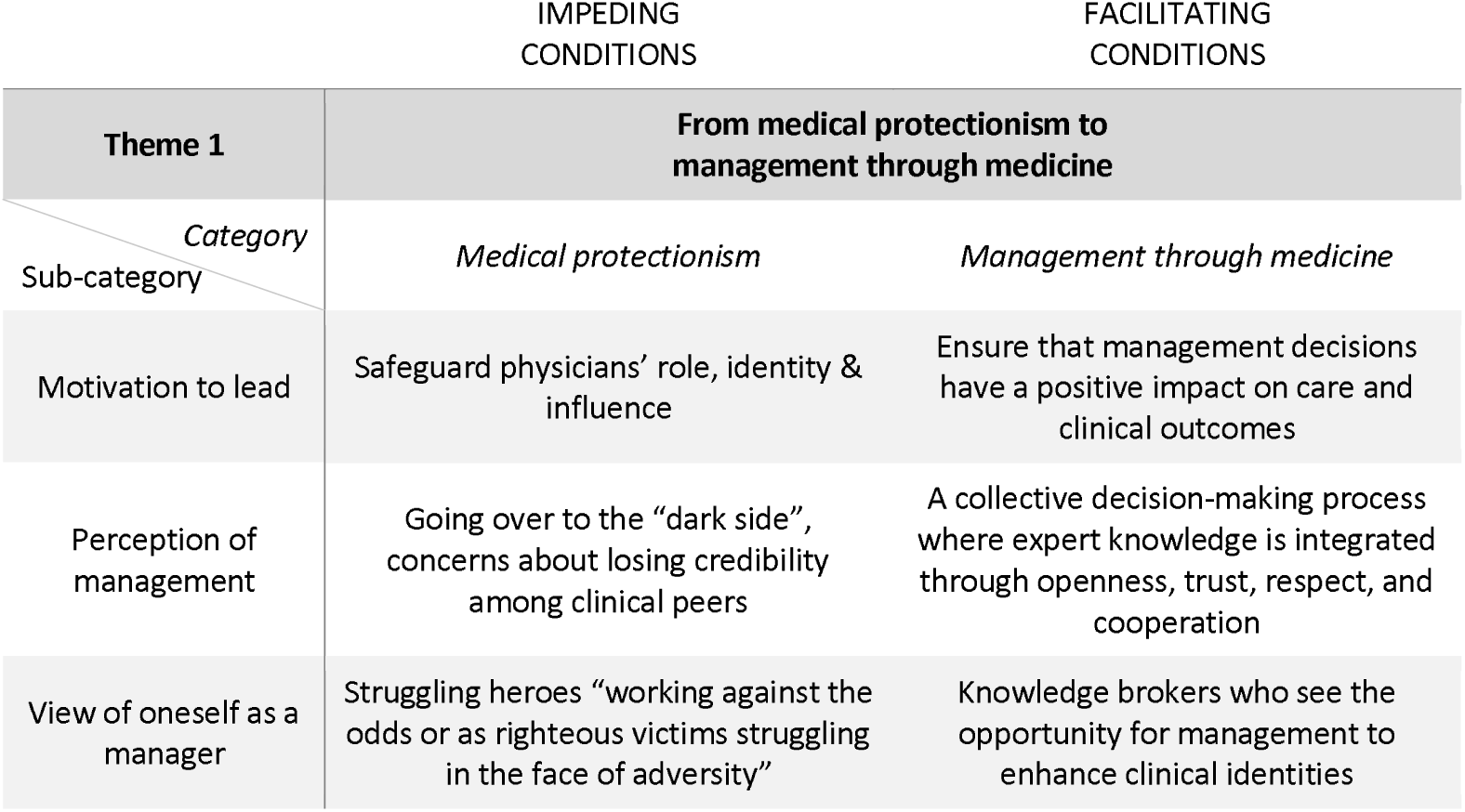
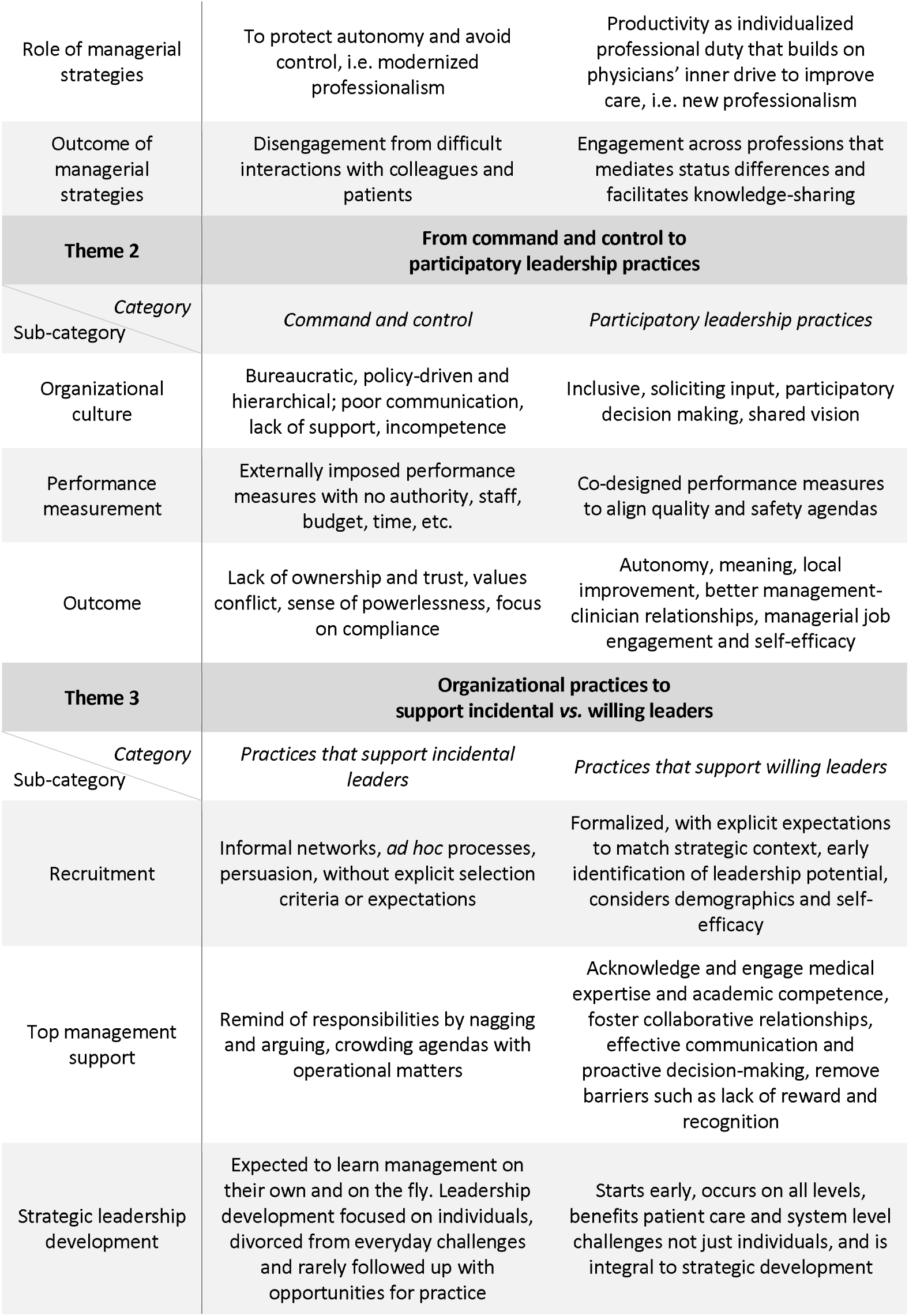
Descriptive themes, categories and sub-categories identified through the thematic synthesis.

|  | IMPEDING<br>CONDITIONS | FACILITATING<br>CONDITIONS |
| --- | --- | --- |
| Theme 1 | From medical protectionism to management through medicine |  |
| Category<br>Sub-category | *Medical protectionism* | *Management through medicine* |
| Motivation to lead | Safeguard physicians' role, identity & influence | Ensure that management decisions have a positive impact on care and clinical outcomes |
| Perception of management | Going over to the "dark side", concerns about losing credibility among clinical peers | A collective decision-making process where expert knowledge is integrated through openness, trust, respect, and cooperation |
| View of oneself as a manager | Struggling heroes "working against the odds or as righteous victims struggling in the face of adversity" | Knowledge brokers who see the opportunity for management to enhance clinical identities |
| Role of managerial strategies | To protect autonomy and avoid control, i.e. modernized professionalism | Productivity as individualized professional duty that builds on physicians' inner drive to improve care, i.e. new professionalism |
| Outcome of managerial strategies | Disengagement from difficult interactions with colleagues and patients | Engagement across professions that mediates status differences and facilitates knowledge-sharing |
| **Theme 2** | **From command and control to participatory leadership practices** |  |
| *Category*<br>Sub-category | *Command and control* | *Participatory leadership practices* |
| Organizational culture | Bureaucratic, policy-driven and hierarchical; poor communication, lack of support, incompetence | Inclusive, soliciting input, participatory decision making, shared vision |
| Performance measurement | Externally imposed performance measures with no authority, staff, budget, time, etc. | Co-designed performance measures to align quality and safety agendas |
| Outcome | Lack of ownership and trust, values conflict, sense of powerlessness, focus on compliance | Autonomy, meaning, local improvement, better management-clinician relationships, managerial job engagement and self-efficacy |
| **Theme 3** | **Organizational practices to support incidental vs. willing leaders** |  |
| *Category*<br>Sub-category | *Practices that support incidental leaders* | *Practices that support willing leaders* |
| Recruitment | Informal networks, *ad hoc* processes, persuasion, without explicit selection criteria or expectations | Formalized, with explicit expectations to match strategic context, early identification of leadership potential, considers demographics and self-efficacy |
| Top management support | Remind of responsibilities by nagging and arguing, crowding agendas with operational matters | Acknowledge and engage medical expertise and academic competence, foster collaborative relationships, effective communication and proactive decision-making, remove barriers such as lack of reward and recognition |
| Strategic leadership development | Expected to learn management on their own and on the fly. Leadership development focused on individuals, divorced from everyday challenges and rarely followed up with opportunities for practice | Starts early, occurs on all levels, benefits patient care and system level challenges not just individuals, and is integral to strategic development |

#### From medical protectionism to management through medicine

Variations in the perceptions of management, views of oneself as a manager, motivation to lead, and the role and outcomes of managerial strategies can be described as medical protectionism or management through medicine.

##### Medical protectionism

Managerial and clinical logics are challenging for physicians to reconcile.[15–18] Medical leaders are perceived to occupy a no-mans-land,[19] often not meeting the expectations and authority vested in them.[20] Many are concerned with losing their credibility among their peers and becoming outsiders,[21] with management referred to as the “dark side”.[15,17,22] They perceive themselves as struggling heroes, “working against the odds or as righteous victims struggling in the face of adversity”.[15]

Physicians’ motivation to be involved in leadership is to safeguard their autonomy, identity, status, influence, and to resist changes tied to their specialty independent of the organization’s needs and goals. They adopt or adapt managerial practices and accept managerial roles as a custodial strategy, referred to as “paradigm freeze”.[6,23–26] This “modernized professionalism” creates new forms of self-regulation and self-management, such as resisting managers’ attempts to control patient safety programs; focusing on minimum necessary reporting; selectively participating in managerial meetings; sending out last minute meeting agendas to limit managers’ participation; or concealing the significance of certain decisions.[24,27] As clinical managers appear to adhere to managerial control, their clinical identity and professional objectives remain unaffected, i.e. loyalty to the profession has trumped loyalty to the organization.[19,24] These dynamics result in personal struggles causing clinicians to disengage from difficult interactions with colleagues and patients, and the medical decision-making suffers.[28] When ignoring as opposed to engaging with these aspects of professional cultures, professional resistance to change can be triggered.[29]

##### Management through medicine

Studies suggest an opportunity to move beyond an adversarial view of management and medicine.[24] Knowledge brokering as described in the concept of hybrid managers can be replaced with an integrative mindset where management is intertwined with expert knowledge through openness, trust, respect, and cooperation, and understood through its impact on clinical practice,[16–18,24,30,31] so that medical leaders can enhance their physician identities by bridging management and medicine.[22]

As physicians are driven by a desire to make a difference, improve, and innovate and want to be engaged and become good leaders,[32,33] managerial discourse should build on their inner drive, resonate with their mental models, and be anchored in quality improvement, i.e. a “professional path”.[18] This “new professionalism” identifies productivity as a route to self-governance where medical leaders achieve superior performance by defining their own and other’s roles, connect staff, and focus on goal attainment.[18,34–37]

Management through medicine has been strengthened by new roles for physicians (e.g. pathway coordinators and hospitalists) and multi-professional, team-based service delivery approaches which mediate status differences and facilitate knowledge-sharing across professions.[16,32,38,39] These allow physicians to enter managerial work earlier in their careers,[20] and thereby improve their managerial capabilities, including building their social capital and developing different perspectives on problems and solutions.[16,18] While some leaders feel it is inappropriate to retain clinical commitments due to a risk of being seen as partisan in relation to a specialty or service,[22] most choose to continue clinical practice to maintain a sense of belonging, enhance legitimacy, and provide inspiration and insights into daily work, but also to keep the option of returning to clinical work in case of failure as a leader.[22,31,33,40]

As discourse has not only a descriptive but also a performative role, there has been a conscious move to replace the managerial discourse with a leadership discourse.[35,40,41] The term “medical leadership” resonates better with professional groups, can remove tensions between operational requirements and visionary aspirations, and potentially influence new work practices.[35,41]

#### From command and control to participatory leadership practices

Organizational attributes, strategies in performance measurement and their outcomes can be described either as management trough command and control or as participatory leadership practices.

##### Management through command and control

Bureaucratic, policy-driven, and hierarchical workplaces with poor organizational communication practices, lack of support for innovation, conflicts, and incompetence hinder physician engagement.[32,42–44] Matrix organizations and distributed leadership are presented as solutions, yet medical leaders still believe that real decision-making power lies outside of care environments, is externalized, and hierarchical.[15,45] Decentralization has been highlighted as a contributor to role ambiguity and overload.[17,46] A lack of support leads physicians to rely on personality, status, and hierarchy, which are insufficient for complex tasks.[28,47] This has a disengaging effect.[32]

Clinicians on different management levels in hospitals and primary care describe a sense of powerlessness over being held accountable for performance measures and organizational issues with neither the authority, staff, budget, time, nor support to actually implement change or to improve.[15,21,32,45,46,48] The overwhelming number of performance targets and guidelines that are externally imposed conflict with professional values and interests,[26,49] and are so demanding that managers tend to focus on compliance, rather than the proactive development of new solutions, and interest in knowledge creation and innovation diminishes.[16,49] Lack of internal peer support makes medical leaders feel that they are alone with their managerial challenges with limited opportunities to discuss and develop ideas for improvement.[21,44] The positive potential of performance measurement, particularly in terms of monitoring quality data, does not materialize due to a lack of ownership over the indicators and also because of problems with access to data and insufficient resources for data collection.[21,48] The time delay between patient safety incidents and quality reports undermine clinicians’ confidence in the data[49] and impede accountability for outcomes.[28]

##### Participatory leadership practices

Physicians need to be given the opportunity to exhibit inclusive leadership behaviors such as explicitly soliciting team input and engaging in participatory decision-making, which in turn help improve their managerial self-efficacy.[3,50] Working with a shared vision, demonstrating compassion, and other positive leadership experiences are associated with managerial job engagement, performance, and participation in leadership activities.[32,44,51–53]

Co-designing performance measures with clinicians, motivates, provides autonomy, makes measurement meaningful, enables local improvement, and can reinforce professionalism in ways that improve the manager-clinician relationship.[24,35,38,39,48,54–56] Physicians can be engaged through continual dialogue to align agendas for quality and safety[21,39,57] and through the design of service delivery.[3,15] Anchoring quality improvement in professional practice, and combining it with education and research, lead to positive views on further improvement initiatives.[3,21,25,29,32,35,38,39,56]

Similarly, budgetary participation supports accountability through autonomy as it positively correlates with budget goal commitment, use of budget information, and therefore budgetary performance.[58] It also improves overall managerial job engagement as it affects managerial self-efficacy, helps to identify with organizational goals, and, along with role clarity, promotes constructive managerial work attitudes.[51,58–60] Tools, such as managerial accounting could co-exist with clinical practice as they are often seen as technical tools without threat to professional autonomy.[24]

#### Organizational practices to nurture willing vs. incidental leaders

Organizational practices that nurture either willing or incidental leaders can be described in terms of recruitment of medical leaders, top management support, and strategic leadership development.

##### Recruitment of medical leaders

Health care organizations require a large number of clinically trained leaders at all levels of the organization, in particular high quality first-line management.[6,19] Despite the fact that interest in leadership can arise from boredom with clinical routine and a desire to take on new challenges,[23] sixty-two percent of executive positions in teaching hospitals are filled by external hires, which suggests a failure to identify, develop, and promote emerging leaders from within the organization.[40,61] Recruitment of medical leaders most often occurs through informal networks and succeeds through the persuasive ability of the current managers, without explicit selection criteria or expectations related to performance objectives, goals, or measures of success.[23,33,44,45] When formal recruitment procedures are followed, the process still tends to be *ad hoc* and lessons learned by search committees are neither captured nor shared. The consequence of these coercive or *ad hoc* approaches that generate “incidental” leaders instead of “willing” leaders can be seen early in leadership development, where the latter are more able to “absorb” or construct managerial expertise.[40,51,62]

To avoid “incidental” medical leaders, recruitment should be formalized, identification of leadership potential should start at an early stage by engaging in conversations with front-line physicians, and these future physician leaders should be supported and molded through opportunities to lead new initiatives.[2,23,32,40,44] In that process, assessment of professionals’ self-efficacy as a predictor of motivation to lead is recommended.[46] Selection of leaders should be part of the overall talent management system[61] and the position should be matched to the strategic, structural, and political context.[21,45,63] Demographics should be considered to avoid management by the “old boys’ club”.[32] The recruitment process should set clear expectations on what is acceptable professional behavior as a medical leader in order to be able to enforce these behaviors in case of a mismatch.[63] While the most frequently displayed and among the most valued leadership attributes among physicians is being inspirational, it has the least impact on staff satisfaction.[4] Those physicians who demonstrate interest in quality, patient safety, and overall leadership aptitude should be sought.[21,45,63] Backgrounds as general internists and practicing hospitalists (or other holistic specializations) seem favorable.[16,21]

##### Top management support

Senior leadership teams, particularly CEOs, manage physicians by nagging, arguing, and reminding them of their responsibilities, i.e. they fail to meaningfully engage medical leaders.[43,64,65] CEOs and senior leadership teams tend to crowd medical leaders’ agendas with numerous committees or “strategic” meetings that are filled with operational, not strategic matters.[21,41,44]

A questionnaire study among staff at the NHS concluded that effective leadership practice (e.g. engaging staff and collaborators in achieving a compelling vision) is correlated with hospital performance.[1] In addition, there is a correlation between how effectively boards work with quality of care and how well executive management teams as a consequence monitor quality and manage operations.[55,57,66] Top-level teams should be stable and acknowledge physicians’ medical expertise and academic competence,[52,65] and foster collaborative relationships, effective communication, diffusion of expert knowledge between managers and professionals, and demonstrate a proactive culture for decision-making.[24,32,49,54,63,67] They also need to remove barriers to medical leadership, e.g. reduce the burden of administrative tasks related to information technology, performance analysis, and financial management; lack of financial incentives; time commitment pressures; overall lack of support, and challenges tied to the timing, location, and process of managerial meetings.[17,20,23,28,31–33,44] This can be done by setting clear expectations[44], introducing collective leadership[19] or through hybrid organizations.[68] The latter resonates well with the idea of professional bureaucracies where staff has greater influence on decision making than people in formal positions of authority.[19]

##### Strategic leadership development

Current undergraduate medical education programs provide only limited opportunities for professional development and neglect strengthening the ethos and professionalism that would make physicians better fit for the purpose of their work.[21] During their clinical careers, they are not sufficiently exposed to professionals who are able to develop their managerial mindset.[20] Management skills are perceived to be in conflict with a medical case-orientation and interventionist professional action.[29] Previous experiences of being a manager at the unit level are not enough either – physicians still have the tendency to be occupied with small scale problem solving which makes it difficult to develop the essential strategic hospital-wide perspective.[20] Even if physicians enter management, they see this merely as an intermediate role.[31] Medical leaders feel they are thrown into their roles and then expected to learn management on their own and on-the-fly.[23,33] Traditional leadership development programs tend to emphasize the difference between management and leadership, which adds to the problem of translating these to practical situations where they actually are intertwined.[41] Leadership training is rarely followed up with concrete opportunities to engage in hospital strategy development.[20]

The introduction of management competencies needs to start early and focus on taking initiative, organizational and system understanding, becoming team players, communication, and shared decision-making.[20,28,65] Leadership development provides four important opportunities to improve quality and efficiency in healthcare, by (1) increasing the caliber of the workforce, (2) enhancing efficiency in the organization’s education and development activities, (3) reducing turnover and related expenses, and (4) focusing organizational attention on specific strategic priorities.[69] Training should improve leaders abilities to address system level challenges and benefit the service, not just the individual.[19,70] Development initiatives create a space for informal conversations that shape attitudes towards teamwork, safety, management and working conditions.[16,41,71] Investments in leadership development should be made at all organizational levels and be seen as part of the strategic development of an organization.[19]

Teaching approaches should move from competency to capability development through integration with ongoing improvement efforts where the focus is on participants’ actual challenges as opposed to merely talking about problem solving.[22,23,29,62,63] Everyday work practices can become opportunities to develop and test new approaches to service provision and to acquire management and leadership skills (e.g. via efficient meetings, medical teamwork, joint decision-making, and the delegation of responsibilities).[25,29] Inter-professional education and training are critical to improve managerial self-efficacy, interest, and readiness to be involved in managerial work.[32,38,40,46] Through mentoring, coaching and networks, medical leaders with similar roles can share experiences, tools, and strategies.[21,22,32,40]

## DISCUSSION

This review provides an in-depth analysis of the conditions instrumental for medical leadership to have an impact on organizational performance. Based on the identified conditions that facilitate or impede the influence of medical leadership, two opposing schemata related to willing *vs*. incidental leadership can be discerned (Figure 4).

**Figure 4.**
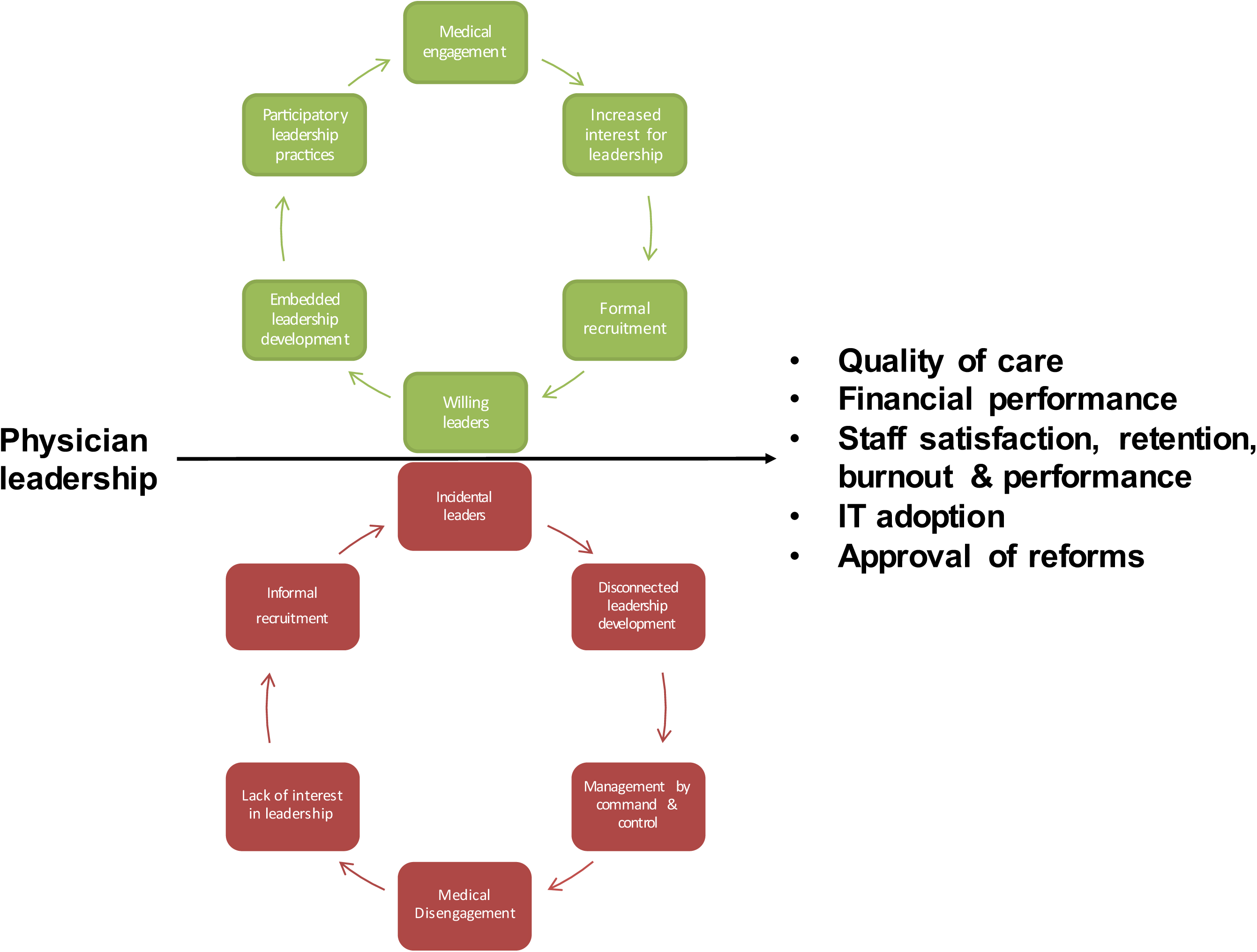
The virtuous and vicious cycles of medical leadership.

The virtuous cycle describes a set of interdependent strategies that help to anchor management in medicine. The pivotal point is to identify willing leaders who are committed to continually improve their own management and leadership competencies. They are nurtured by an embedded leadership development strategy that fosters participatory leadership practices. Participation cultivates medical engagement among staff and thereby increases interest in leadership roles and management positions. This, in turn, contributes to favorable conditions for formal recruitment and expands the recruitment pool of future willing leaders.

In the vicious cycle, managerial positions are filled by incidental leaders with little interest to improve their own leadership competencies. The lack of interest is reinforced by disconnected leadership development efforts that are perceived as irrelevant to the improvement of health care. Managers mimic historically dominant managerial approaches, i.e. management by command and control, which leads to medical disengagement among staff. Disinterest in leadership roles encourages informal recruitment practices which perpetuates the risk for incidental leaders.

The findings of this review resonate with the emerging field of research tied to physician or medical engagement. Medical engagement is defined as a reciprocal relationship between the individuals and the organizational system: “the active and positive contribution of doctors, within their normal working roles, to maintaining and enhancing the performance of the organization, which itself recognizes this commitment, in supporting and encouraging high quality care”.[52] A recent review elaborates that physician engagement is about “regular participation of physicians in (1) deciding how their work is done, (2) making suggestions for improvement, (3) goal setting, (4) planning, and (5) monitoring of their performance in activities targeted at the micro (patient), meso (organization), and/or macro (health system) levels.”[72]

While Spurgeon *et al*.[63] ask if it is medical leadership or medical engagement that is needed for better performance, we suggest that medical engagement is intimately dependent on the quality of medical leadership. The virtuous cycle of medical leadership illustrates how medical leadership can intervene at the individual, organizational and system levels to enhance medical engagement. At the individual level, medical leaders can explicitly use their medical knowledge to interpret and explain the medical consequences of managerial decisions.[73] This would demonstrate commitment to improve health care, model an integrative view of management and medicine, and subsequently, enhance professional identities. At the organizational level, medical leaders should formalize recruitment processes, get top management teams to acknowledge and engage medical expertise and academic competence, and embed leadership development in medical practice through quality improvement. Finally, the highest level of medical leadership, including political decision makers, need to develop an inclusive and collaborative culture characterized by openness, trust, and respect, by engaging health professionals in the design and monitoring of performance measures. These combined efforts will not only cultivate medical engagement and by that improve the performance of individual health care organizations. They will also enable a shift to new leadership paradigms suitable to the complexity of health care,[74] and establish conditions favorable for large-system transformation and health care reform.[75]

In terms of future research, the field of medical leadership would benefit from studies conducted in primary care, include leaders at other than senior managerial levels, and from non-Anglo-American settings. While we came across a few studies on gender balance and internationalization of the clinical workforce, perspectives on the consequences for medical leadership are lacking. Qualitative studies could further deepen our understanding of the relationship between management and medicine in everyday clinical practice in order to inform leadership development and human resource management efforts. Finally, this review alludes to a need to design and evaluate medical leadership development programs that are theory-based, evidence-informed, and organizationally embedded.

This review is limited by the quality and heterogeneity of included studies. Quality appraisal of the individual studies in terms of strength of evidence was not conducted due to the reviews broad focus which lead to significant diversity of research designs. Since the search was timebound to capture contemporary evidence and limited to three databases, we cannot guarantee that all relevant articles were found. While plausible correlations between conditions and performance outcomes are explored, to establish causality requires other approaches to test and determine the strength of the relationships.

## CONCLUSION

The identification of the virtuous or vicious cycles of medical leadership can help us better understand how medical leadership can be both a boon or a barrier to the positive impact that health care organizations desire for their patients, staff, and society. We can choose to either create willing leaders through medical engagement or accept incidental leaders through medical protectionism. This complex challenge involves questioning conventional wisdom on management and medicine in favor of more participative practices that require long-term investments at the individual, organizational, and system levels.

## Data Availability

Data for this study includes peer-reviewed empirical studies and literature reviews. The detailed overview of included studies is provided as supplementary information. Data extraction and analysis was performed in NVivo qualitative data analysis software; QSR International Pty Ltd. Version 10, 2012. The NVivo file can be made available upon reasonable request.

## Authors’ contributions

MS, CS, MB, and PM designed the study. MS conducted the search with support from a professional research librarian. MS screened titles, key words, and abstracts for inclusion. All authors screened full texts for inclusion. MS extracted data and performed line-by-line coding of the included studies. Based on codes, all authors collectively developed descriptive and analytic themes. MS drafted the manuscript. All authors read, revised, contributed to, and approved the final manuscript. PM was the principal investigator.

## Acknowledgements

The authors wish to thank the Karolinska Institutet University Library for their help with the search strategy and access to articles, and the Swedish Medical Association for their continued inquiries into physician leadership.

## Ethical approval

An ethical vetting was deemed unnecessary.

